# Speedy and satisfying: Real-time Location System increases Emergency Department efficiency and decreases frustration with finding medical equipment

**DOI:** 10.1101/2023.11.20.23298180

**Authors:** Sandhya LoGalbo, Sophia Trojanowski, Alexandria Slusser, Lisa Iyeke, Lindsay Jordan, Mark Richman

## Abstract

**INTRODUCTION:** We evaluated the impact of a real-time locating system on emergency department efficiency and frustration locating mobile otoscope/ophthalmoscope carts.

**DATA AND METHODS:** Thirty ED providers were selected on-shift, each serving as their own control. Investigators hid two mobile otoscope/ophthalmoscopes carts (with and without the RTLS device) equidistant from the center of a provider’s work area. Providers were timed finding both devices and were queried regarding feelings about the search experience.

**RESULTS:** RTLS was associated with statistically-significantly less time locating equipment (average of 25 vs. 92 seconds) and percent of providers requiring 30, 60, 90, and 120 seconds to find the device. Providers felt finding the mobile cart with RTLS was easier; all rated finding the RTLS-tagged cart as easy. Without RTLS, two-thirds of subjects reported either frustration or extreme frustration vs. 3% with RTLS. All differences in comparisons of subjective experience were statistically-significant. Annual time and cost saving with RTLS would be 116.4 hours ($9,135.66).

**CONCLUSION:** RTLS in EDs can decrease time and frustration associated with finding equipment and is cost-effective. Frustration is a common driver behind burnout in Emergency Medicine. Use of RTLS technology might improve the provider experience and, thereby, reduce burnout levels.

## INTRODUCTION

Real-time Location Systems (RTLS) are electronic tracking services that use radio frequencies (based on global positioning systems (GPS)) that are attached to items of interest or value and used to locate them. Use of RTLS is associated with increased efficiency of finding objects and persons. In healthcare RTLS has been utilized to monitor patients, medical staff, and medical equipment to maximize patient care and increase hospital productivity in various clinical settings, including Emergency Departments (EDs) and surgical wards.^1,2^ The feasibility and practicality of RTLS in healthcare settings is dictated by the energy consumption, detection range, size, and accuracy of the RTLS device. The innovation of bluetooth technology and wireless options allowed creation of new, affordable, and functional devices compared with previous systems. The main types of RTLS are RFID and bluetooth. Radio-frequency identification (RFID) RTLS,^3^ which was the original platform used for this function, improved patient care but had high costs of radio wave-compatible infrastructure, and technical problems with chips compared with bluetooth technology RTLS.^4^ Bluetooth RTLS has excellent Received Signal Strength Indication (RSSI), cellular device compatibility, and large connection and identification range. One such bluetooth RTLS device is the “Tile®” tracker,^5^ which can potentially save time and effort finding equipment in busy settings such as the ED.

While the volume of ED patients continues to increase,^6^ the number of EDs is stable or declining.^7^ Emergency Physicians are required to efficiently provide optimal patient care within a reasonable amount of time. Difficulty accessing functional equipment can endanger patients and cause frustration, poor patient experiences, and provider frustration and burnout.

Otoscopes and ophthalmoscopes are used daily by ED providers to examine patients. The hospital surveyed in this study uses otoscopes and ophthalmoscopes attached to rolling carts. The ED has 4 carts, but 7 treatment areas and 47 treatment rooms. Carts are often left by providers in the rooms, either because they forget them there or because it is inconvenient to bring the device back to a common placement area, or, at least, a hallway, where they can more-easily be found. Inability to easily-find a mobile otoscope/ophthalmoscope device has been a long-time inconvenience for providers, who spend valuable time locating these devices. High ED volumes and hectic environment lead to an increase in entropy,^8^ made worse by providers continually searching for otoscopes/ophthalmoscopes. This barrier to optimal patient care is a source of frustration to providers.

This study aimed to determine if the Tile® device and app increases the efficiency of locating the mobile otoscope/ophthalmoscope cart by providers, as well as the impact, if any, on frustration levels in locating these devices. We hypothesized that adding a Tile® tracking device to mobile otoscope/ophthalmoscope carts would decrease the time and frustration associated with finding such equipment.

## DATA AND METHODS

We performed a randomized, controlled trial in which each adult ED subject acted as their own control. The subjects were Attending physicians, Residents, Physician Assistants, and Nurse Practitioners. Staff searched the ED for the otoscope/ophthalmoscope carts with and without the Tile^®^ device. The 30 subjects were selected through a convenience sample and all participants were asked for their consent and availability on-shift. We performed this study while the providers were on shift, mimicking what would happen in true circumstances.

Investigators hid two mobile otoscope/ophthalmoscopes carts, one with a Tile® device and one without the Tile® device. Both carts were equidistant from the center of the zone (eg, Red Zone, Green Zone) where the provider subject was stationed. A provider subject from that zone was asked to find both devices: first the one without the Tile® device, then the one with the Tile® device.

We created a questionnaire survey to capture objective and subjective data regarding the otoscope/ophthalmoscope search experience.

1. Objective questions: Investigators recorded the time (in seconds) it took the provider subject to find the mobile otoscope/ophthalmoscope cart with and without the Tile® device.
2. Subjective questions: Provider subjects answered questions regarding their feelings about the experience searching for the mobile otoscope/ophthalmoscope cart. After completing both trials, subjects took a 13-question paper survey detailing their demographics, experience (convenience), and emotions. Emotions were ranked using a Likert scale from 1 to 5.

Sample size: Based on power = 80%, alpha = 0.05, and effect size 75% (from formative research), it was estimated the study required 17 subjects. Since subjects served as their own control, a total of 30 participants were enrolled to ensure statistical reliability,^9^ each of whom sought both the Tile® and non-Tile® devices. Continuous variables were compared via a means t-test while categorical variables were compared with a Chi-Square test. Statistical significance was set a priori at p <0.005. Data was entered into a REDCap® database and analyzed utilizing Microsoft Excel® 2017.

As a quality improvement project, this study was deemed not to meet the definition of research by the Institutional Review Board in its Human Subjects Determination Request evaluation.

## RESULTS

Use of the Tile® device was associated with less time locating a mobile otoscope/ophthalmoscope cart (average of 25 vs. 92 seconds) (**Figure 1**). Only 13.3% of providers found the mobile cart without the Tile® device in under 30 seconds, while 73.3% found the cart in under 30 seconds when it had an attached Tile® device (**Figure 2**). Similar decreases were found when comparing rates of cart discovery after 60 seconds. No provider took more than 90 seconds or more to find a Tile®-tagged cart; without the Tile® tag, nearly 45% took at least 90 seconds and 35% took 120 seconds (**Figure 3**). Providers felt that finding the mobile cart without the Tile® device was 67.5% more difficult vs. finding the Tile®-tagged cart, and 33.33% described finding the non-tagged cart extremely difficult (**Figure 4**). In contrast, all subjects rated finding the Tile®-tagged cart as either very easy (70%) or slightly easy (30%) (**Figure 5**). Without RTLS, two-thirds of subjects reported either frustration or extreme frustration (**Figure 6**). With a tracking device, only 3% reported frustration or extreme frustration (**Figure 7**).

**Figure 1.**
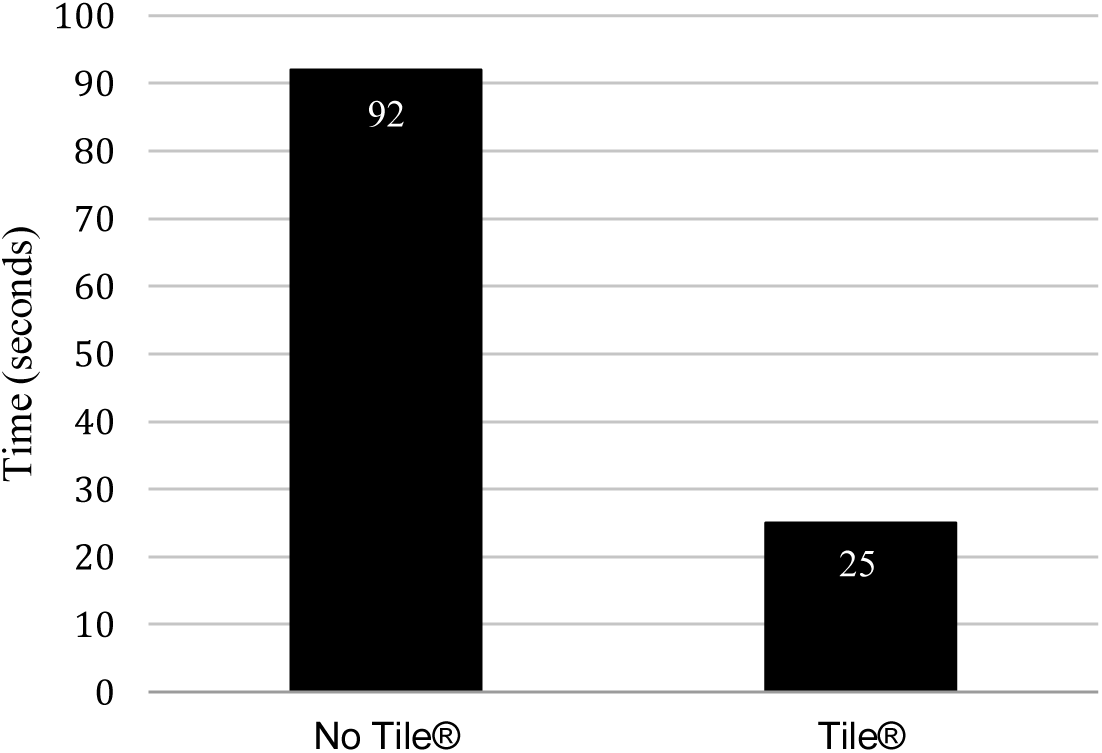
Average time in seconds to find an otoscope/ophthalmoscope cart with and without Tile® device.

**Figure 2.**
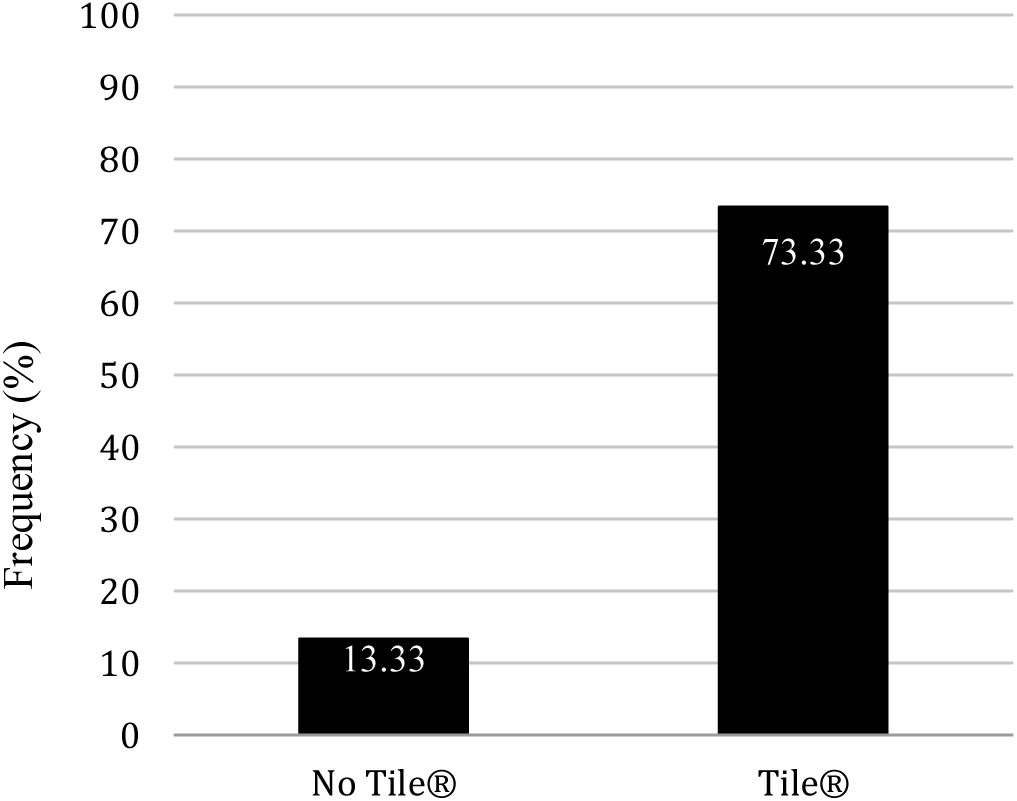
Percentage of subjects who found an otoscope/ophthalmoscope cart with and without a Tile® device in under 30 seconds.

**Figure 3.**
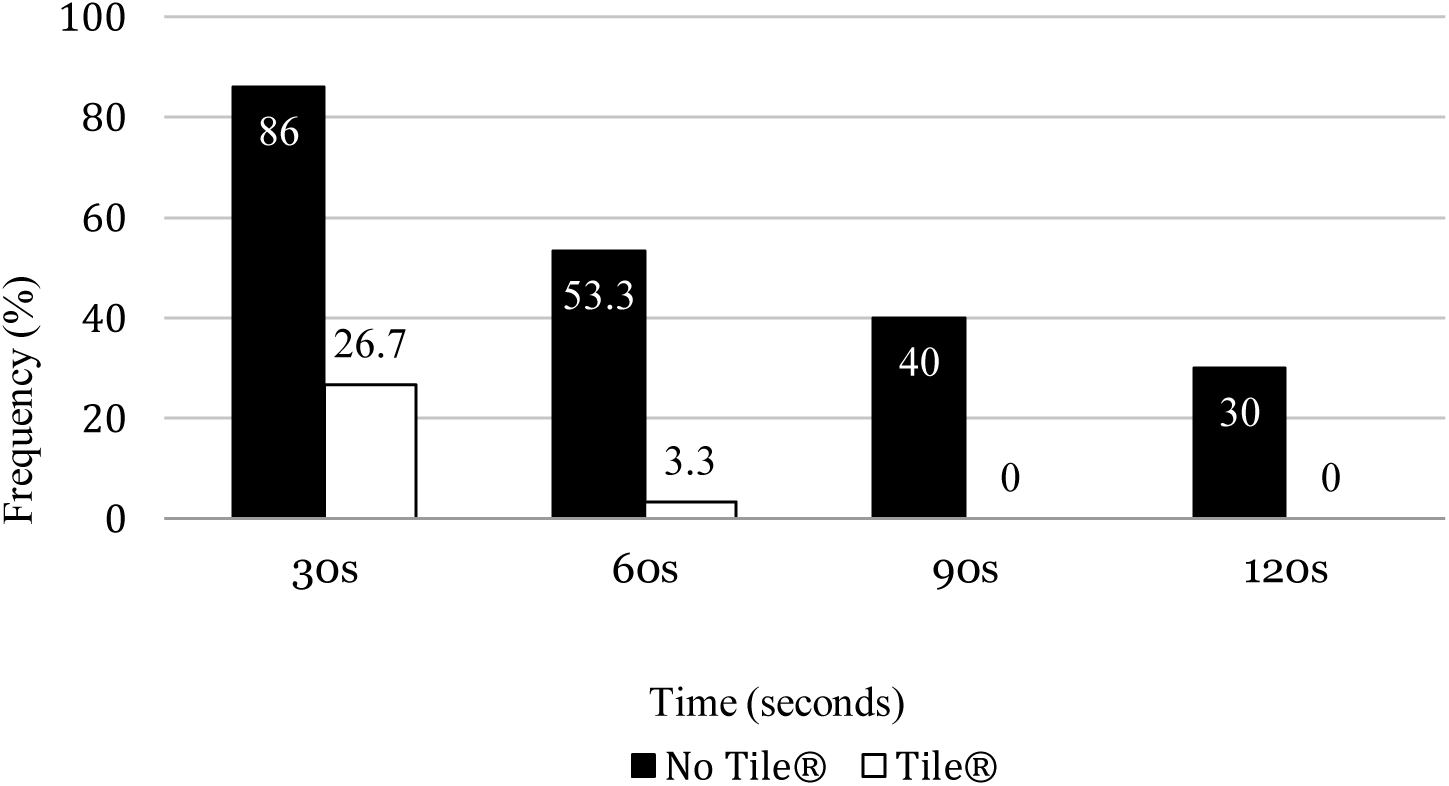
Percentage of subjects who found an otoscope/ophthalmoscope cart with and without a Tile® device in over 30 seconds, over 60 seconds, over 90 seconds, and over 120 seconds respectively.

**Figure 4.**
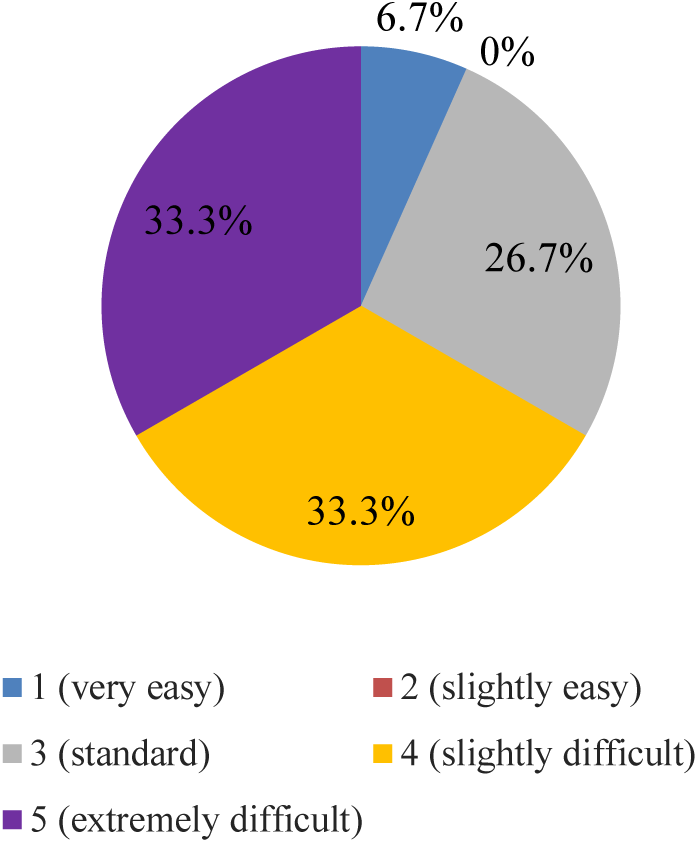
Subjective ratings of ease of locating a mobile otoscope/ophthalmoscope without a Tile® device.

**Figure 5.**
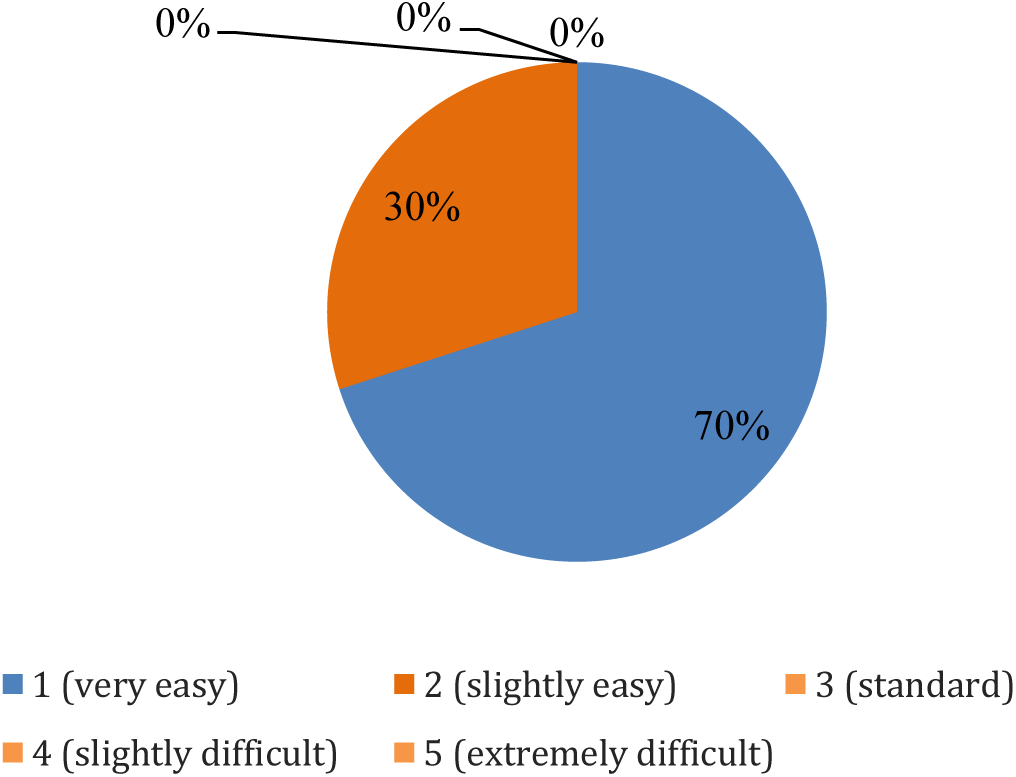
Subjective ratings of ease of locating a mobile otoscope/ophthalmoscope with a Tile® device.

**Figure 6.**
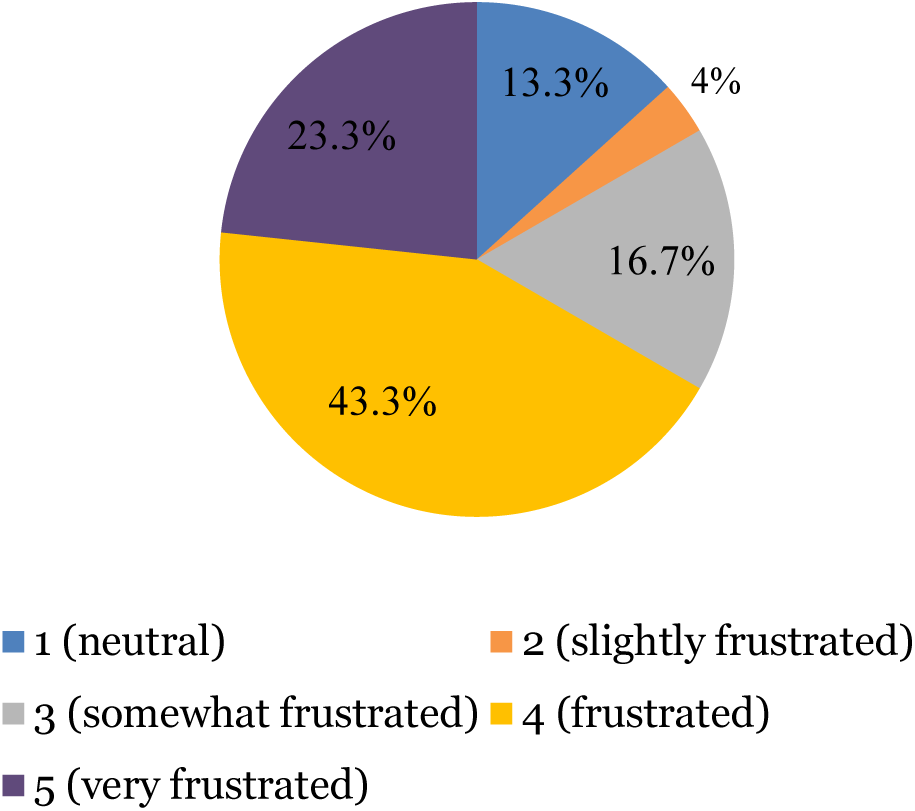
Subjective ratings of frustration while finding a mobile otoscope/ophthalmoscope cart without a Tile® device.

**Figure 7.**
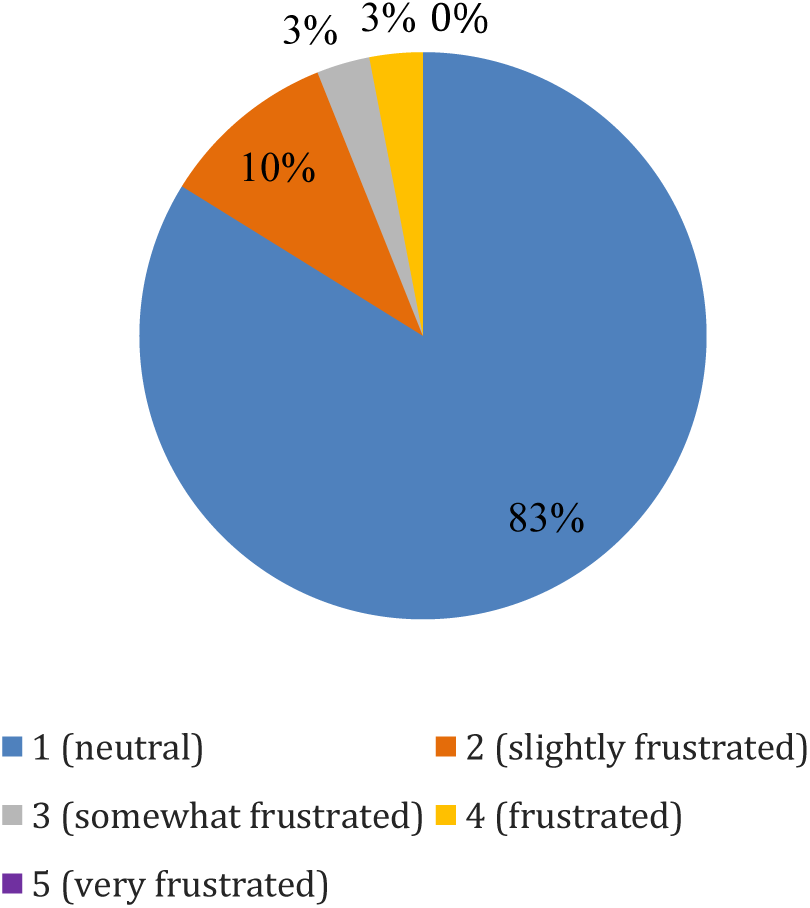
Subjective ratings of frustration while finding a mobile otoscope/ophthalmoscope cart with Tile® device.

Providers stated that, on average, they use the otoscope 2 times/week, for a total usage of 100 times per year. Given the estimated annual time currently spent searching for the otoscope/ophthalmoscope carts without the Tile® device, and the estimated time saving with the Tile® device, it is estimated that using the Tile® device would save 116.4 hours annually and a cost of $9,135.66 ($12,575.07-$3,439.41) (**Table 1**, **Figures 8** **and** **9**).

**Figure 8.**
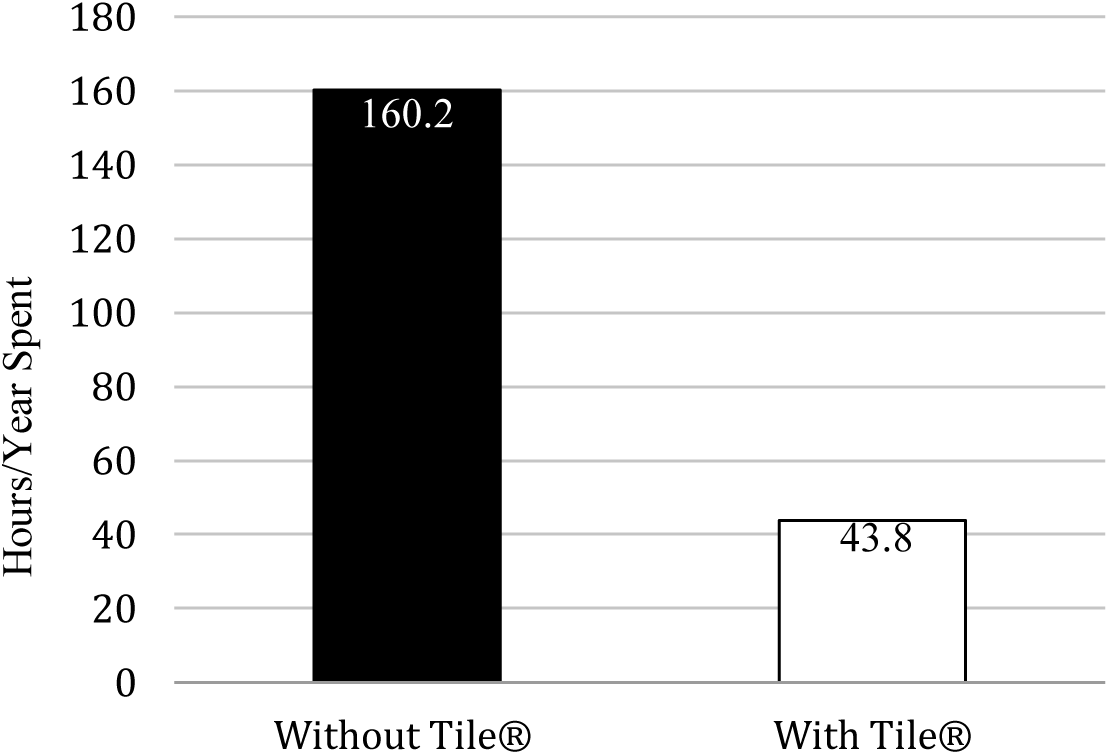
Average hours/year spent locating otoscopes/ophthalmoscopes without vs. with the Tile® device.

**Figure 9.**
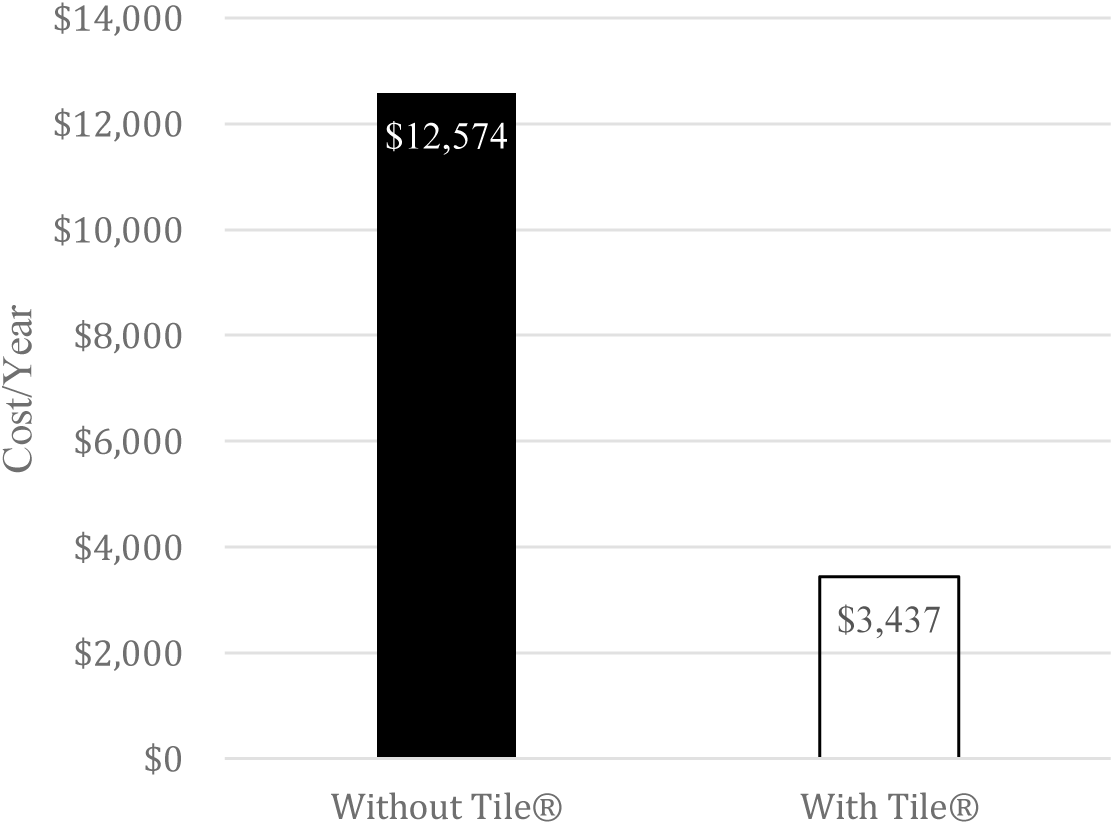
Average cost of time spent locating otoscopes/ophthalmoscopes over the course of a year without vs. with use of a Tile® tracking device.

**Table 1.** Annual time and cost savings searching for mobile otoscope/ophthalmoscope with vs. without Tile® device.

| Type of provider | Hourly pay of provider | Annual # of hours searching for otoscope/opthalmoscope carts without the Tile® | Annual # of hours searching for otoscope/opthalmoscope carts with Tile® | Annual cost searching for otoscope/ophthalmoscope carts without the Tile® | Annual cost searching for otoscope/ophthalmoscope carts with the Tile® |
| --- | --- | --- | --- | --- | --- |
| Physician Assistant | \$64.10 | 80.1 | 21.9 | \$5,120 | \$1,403.79 |
| Resident Physician | \$25.60 | 40.05 | 10.95 | \$1,036.80 | \$280.77 |
| Attending Physician | \$160.26 | 40.05 | 10.95 | \$6,418.27 | \$1,754.85 |
| **Total** | | **160.2** | **43.8** | **\$12,575.07** | **\$3,439.41** |

## DISCUSSION

Delays in accessing medical equipment may interfere with patient care and productivity and contribute to poor patient experiences and healthcare professional burn out. Implementation of real time location systems (eg, the Tile^®^ device) on mobile otoscope/ophthalmoscope carts substantially decreased the time to find the devices, improving provider productivity and decreasing frustration.

Utilizing calculations of mean time of finding the mobile otoscope/ophthalmoscope with and without the Tile® device and average salaries for ED staff, it was found that this hospital would save $3,716 per Physician Assistant, $756 per Resident, and $4,663 per Attending Physician, respectively, per year, in time spent searching for the otoscope/ophthalmoscope devices.

Other healthcare facilities have incorporated and utilized RTLS to locate medical devices efficiently. Atrium Health Wake Forest Baptist Medical Center produced a five-year roadmap of RTLS, tagging over 17,000 pieces of mobile medical equipment to address service excellence, patient safety and satisfaction, and operational excellence and efficiency, and transformed healthcare delivery (SPOT). The hospital system has saved approximately $3.5 million dollars thus far and reduced patient wait times. Similarly, an infectious disease outbreak epidemiology simulation exercise in Singapore’s Sengkang General Hospital identified three times the number of contacts using contact tracing via patient- and staff-tagged RTLS compared with tracing via EHR query, reducing by ∼97% the time and staffing required to conduct the tracing.^10^ The NIH published a series of case studies on how RTLS impacted hospital systems. Christiana (Newark, DE) health system utilized RTLS software within infrared tags to track patient movement in real time, leading to a 5% decrease in turnaround time in the ED (despite an increase in volume of over 7% during flu season), a decrease by 24% in rates of patients leaving without being seen, and an increase in patient satisfaction due to improved ED flow. Likewise, Providence Holy Cross Medical Center (Los Angeles, CA) adopted a patient tracking system to adapt to a growing volume of patients (occupancy rates 80-110%). Implementation was associated with an 11.5% increase in inpatient admissions, 10.6% decrease in average length of stay, 16.3% increase in ED visits, and 25.3% decrease in number of patients who left without being seen. Overall, the vast majority (95%) of organizations (ranging from 25 beds to large integrated delivery networks) using RTLS cite operational efficiency gains.^11^ This was corroborated by a systematic review done by Overmann et al.^2^ which found RTLS improved workflow and patient care.

## LIMITATIONS

### Technical and financial limitations

Using a shared RTLS device account, there is no limit to the number of phones to which a particular RTLS tag on a particular device can transmit radiofrequency signals. However, occasionally, the Tile^®^ phone app would not connect (or would disconnect) from the Tile^®^-tag attached to the otoscope/ophthalmoscope cart due to WiFi issues. Each RTLS device will have a limited radius of communication between the tag emitting bluetooth radiofrequency and the receiving device (eg, smart phone) on which the RTLS app resides. This limits the distance beyond which the tagged device can be reliably found. Likewise, thick walls, as are commonly-found in hospital settings, may prevent barriers between connecting the tagged device to the phone-based app. Finally, although the cost per RTLS tag is modest, should an organization wish to tag a great number of items, the initial cost could be quite high. Even though RTLS can be a time-efficient choice, depending on the department budget, RTLS devices may be an inefficient use of funds compared to more dire necessities in the department.^12^

### Study limitations

This was a single-site study based on a simulation exercise and conducted in a specific area based on convenience and accessibility. Only a small, randomly-selected sample (30) was used, and a majority of the participants were resident physicians. Providers were chosen based on availability at time of the study. This could bias the results, even though the population is consistent with the majority of the population that looks for otoscopes/ophthalmoscopes. Additionally, providers searched a designated zone, not the entirety of the ED, because of the bluetooth radius of the Tile^®^ device. RTLS may have better performance in simulation, as, in “real-life” situations, mobile carts are likely to move beyond the limited bluetooth radius to which we constrained the carts during this study.

## CONCLUSION

Despite limitations, the real-time location system (specifically, the Tile® device and app) was easy to use, did not interfere with other medical equipment, reduced provider workload, and was easily-integrated into the ED through a shared Tile® device account. Subjective surveys indicated the technology was highly-accessible and readily-understood. Additionally, a notable decrease in entropy^8^ was seen throughout the ED, reducing stressors that previously disrupted and delayed the transmission of care.

## Data Availability

All data produced in the present work are contained in the manuscript.

## Author Contributions

Mark Richman, Lisa Iyeke, Lindsay Jordan: Conceived of the project and did the initial manuscript draft and literature search.

Sandhya LoGalbo, Sophia Trojanowski, Alexandria Slusser: Created data collection tool, performed data collection and entry, integrated those findings and references into the manuscript, reviewed and edited final manuscript.

## Patient’s Consent

N/A

## Trial/Systematic Review Registry

N/A

## Acknowledgements

None.

## CONFLICT OF INTERESTS

N/A

## FUNDING SUPPORT

N/A

## AVAILABILITY OF DATA AND MATERIALS

N/A

## Notes

### Competing Interest Statement

The authors have declared no competing interest.

### Funding Statement

This study did not receive any funding.

### Author Declarations

As a quality improvement project, this study was deemed not to meet the definition of research by the Institutional Review Board in its Human Subjects Determination Request evaluation. This study was reviewed by the Northwell IRB, which determined it was a performance improvement project and not human subjects research (Human Subjects Research Determination: 23-0100).

## REFERENCES

1 Gholamhosseini L, Sadoughi F, Safaei A. Hospital real-time location system (A practical approach in healthcare): A narrative review article. Iranian Journal of Public Health. 2019; 48(4):593–602. https://www.ncbi.nlm.nih.gov/pmc/articles/PMC6500521/

2 Overmann KM, Wu DTY, Xu CT, Bindhu SS, Barrick L. Real-time locating systems to improve healthcare delivery: A systematic review. Journal of the American Medical Informatics Association : JAMIA. 2021; 28(6):1308–1317. 10.1093/jamia/ocab026

3 Pancham J, Millham R, Fong SJ. Evaluation of real time location system technologies in the health care sector. 2017 17th International Conference on Computational Science and Its Applications (ICCSA). 2017; 1–7. 10.1109/ICCSA.2017.7999645.

4 Yazici HJ. An exploratory analysis of hospital perspectives on real time information requirements and perceived benefits of RFID technology for future adoption. International Journal of Information Management. 2014; 34:603–621. 10.1016/j.ijinfomgt.2014.04.010.

5 Tile by Life360 [Internet]. Tile E-Commerce. 2023 [cited 2023]. Available from: https://www.tile.com/products/deals?utm_campaign=%28P%3AG%29+%28CT%3ABRAND%29+%28GEO%3AUS%29+%28CA%3ABRAND%29_13870698657&utm_source=google&utm_medium=cpc&utm_content=592930507905&utm_term=tile+tracking+device-p&adgroup=130386752611&gclsrc=aw.ds&gclid=CjwKCAiAjPyfBhBMEiwAB2CCImrd_P0nHgUi4tD22Peo0XtMqW7iz1rZtrO5D_auJmI5qS4L6dNXZRoCsnsQAvD_BwE

6 Table EDAd. Emergency department visits within the past 12 months among adults aged 18 and over, by selected characteristics: United States, selected years 1997–2019. Centers for Disease Control and Prevention. 2021. https://www.cdc.gov/nchs/data/hus/2020-2021/EdAd.pdf.

7 Bobb MR, Ahmed A, Van Heukelom P, Tranter R, et al. Key High-efficiency Practices of Emergency Department Providers: A Mixed-methods Study. Academic Emergency Medicine: A Global Journal of Emergency Care 2018. 25(7):795–803. 10.1111/acem.13361.

8 Pendrill L, Espinoza A, Wadman J, Nilsask F, et al. Reducing search times and entropy in hospital emergency departments with real-time location systems. IISE Transactions on Healthcare Systems Engineering 2021. 11(4): 305–315. 10.1080/24725579.2021.1881660

9 Ganti A. Investopedia. Central Limit Theorem (CLT): Definition and Key Characteristics. [Internet]. 2023 [cited 2023]. Available from: https://www.investopedia.com/terms/c/central_limit_theorem.asp#:~:text=A%20sample%20size%20of%2030,representative%20of%20your%20population%20set.

10 Ng GY, Ong BC. Contact tracing using real-time location system (RTLS): a simulation exercise in a tertiary hospital in Singapore. BMJ Open. 2022; 12(10). 10.1136/bmjopen-2021-057522

11 Drazen E, Rhoads J. Using tracking tools to improve patient flow in hospitals. National Library of Medicine. 2011. http://resource.nlm.nih.gov/101567187

12 Real-Time Location Systems (RTLS): Maximizing the ROI. Klas Research. 2011. https://klasresearch.com/report/real-time-location-systems-rtls-2011-maximizing-the-roi/693

